# Physical Activity Phenotypes in Endometriosis Using Unsupervised Learning via Functional Mixture Models

**DOI:** 10.1101/2025.02.10.25322020

**Authors:** Bryan T. Tricoche, Billy A. Caceres, Leslee Shaw, Carol Ewing Garber, Stefan Konigorski, Sahiti Kolli, Thomas Fuchs, Ipek Ensari

## Abstract

**Introduction:** Endometriosis is a chronic condition associated with severe pelvic pain, dysmenorrhea, infertility, and worsening quality of life. Regular physical activity (PA) is effective for pain management and reducing chronic disease symptoms, yet individuals with endometriosis are more likely to be insufficiently active. This study aims to investigate latent profiles of daily PA trajectories in this population via clustering.

**Methods:** The study sample included 173 adults (4,895 person-level days) with a confirmed diagnosis of endometriosis enrolled in the *All of Us Research Program*. PA data was collected from participants using Fitbit wrist-worn trackers. We used 30 consecutive days of data from each individual, allowing up to 10 days of missingness (which were imputed using multiple imputed chained equations). Functional mixture models (FMMs) identified latent PA trajectory clusters using daily step counts as the clustering variable. The optimal number of clusters was determined using the Bayesian information criterion (BIC).

**Results:** The best-fitting FMM identified K=4 distinct clusters (BIC_K=4_ = –11884.442, BIC_K=3_ = –11892.236, BIC_K=5_ = –11907.917). The *High Active* phenotype exhibited the highest volume and variability of step counts over the sample period (Mean = 12777.0, SD=5248.2) as well as the highest volume of moderate-to-vigorous PA (MVPA) minutes (Mean = 70.8). The *High Moderate* phenotype exhibited the second highest volume and variability of step counts (Mean = 9125.4, SD = 3631.0) and second highest MVPA minutes. The *Low Moderate* phenotype exhibited the second lowest volume and variability of step counts (Mean = 6067.5, SD =2797.0) and the second lowest MVPA minutes (Mean = 24.3). The *Insufficiently Active* cluster exhibited the lowest volume and variability of step counts (Mean = 4235.7, SD = 2174.7) and the lowest MVPA (Mean = 16.1).

**Conclusion:** This is the first study to investigate and report distinct PA profiles among a national sample of individuals living with endometriosis, and via objectively estimated PA data. Identifying distinct clusters of PA patterns in endometriosis based on within– and between individual PA variance may help identify those at risk and inform the development of personalized interventions to promote PA and improve overall health outcomes.

## Introduction

Endometriosis is a chronic, estrogen-mediated, inflammatory condition characterized by the growth of endometrial-like tissue outside the uterus, leading to the formation of adhesions and lesions. This condition affects approximately 10% of women of reproductive age and is associated with severe pelvic pain, fatigue, dysmenorrhea, and infertility, significantly reducing the quality of life (QoL) of affected individuals.^1,2^

Regular physical activity (PA) has been recognized as a crucial factor in maintaining overall health and reducing the risk of non-communicable diseases. While engaging in regular, sufficient PA might be impeded by an individual’s chronic disease (e.g., endometriosis pain), PA has been demonstrated to be beneficial for managing endometriosis symptoms, including pain reduction and improved QoL.^3,4,5,6^ Existing studies report that women with endometriosis have lower overall PA volume compared to women without the condition.^3^ This is concerning because regular PA has been associated with reduced risk of developing endometriosis, and can aid in endometriosis symptom management.^4^ A cross-sectional study^7^ investigating the relationship between PA and endometriosis symptoms found evidence that women with endometriosis who engaged in more PA reported lower pain levels and better quality of life. Awad et al.^8^ examined PA patterns in women with chronic pelvic pain, including those with endometriosis, and reported lower PA levels compared to healthy controls. Finally, in a study among individuals with endometriosis, PA was associated with reduced pain severity and improved quality of life.^5^

Existing cohort-based studies in people living with endometriosis mostly rely on self-reported measures, cross-sectional sampling, or tools that are not comprehensive or validated for assessing PA.^9^ Self-reported measures can be subject to recall bias and other sources of measurement error,^10^ therefore limiting the quality of the data and the inferences made from them. Objectively-estimated longitudinal data toward this end can provide a more accurate and comprehensive picture of PA patterns in this population, as well as longitudinal patterns over time.^11,12^ These data subsequently can help clinicians identify those who don’t meet PA guidelines and those who can benefit from tailored PA interventions.

Traditional approaches to analyzing PA data typically involve aggregating the multiple data points per person to get a summary score (e.g., daily or weekly PA volume). This approach, however, does not capture the temporal trends and patterns over time, which could provide additional valuable information about the population of study.^12^ Among individuals with endometriosis, characterizing the temporal and between-individual variations in PA patterns can help identify potentially distinct profiles of PA patterns and the variability of these patterns over time. These “phenotypes” can then be used to tailor PA recommendations and support strategies to each patient’s specific activity patterns and needs. This personalized approach may lead to more effective management of endometriosis symptoms and improved overall health outcomes. In addition, consideration of temporal patterns can help assess the effectiveness of PA interventions by examining changes in activity profiles over time, as well as serve as early indicators of symptom flare-ups or disease progression.

The present study investigates latent profiles of daily PA trajectories among individuals with endometriosis using functional mixture models (FMMs). FMMs consider entire data curves as the unit of analysis, as opposed to discrete data points, making them more flexible and robust for examining the shape of the PA trajectory over time. Accordingly, the primary aim of this study is to identify potential “phenotypes” of PA patterns among individuals living with endometriosis. We use the *All of Us Research Program* cohort to conduct the analyses, which provides a national sample of participants from around the United States (US). Given the literature on the symptomatic heterogeneity in this population, we hypothesized that there would be at least 3 distinct, latent clusters (i.e., “phenotypes”) that correspond to low, moderate, and high activity patterns.^13^ We expected that the identified phenotypes would significantly differ based on temporal fluctuations in PA over time, with higher volumes of PA being associated with greater variability.

## Methods

### Study Sample

The study sample included adults with a confirmed diagnosis of endometriosis enrolled in the NIH *All of Us Research Program*. A detailed description of the research program has been published elsewhere.^14,15^ We used the All of Us Registered Tier Dataset v6, which was made available in June 2022, for the analysis.^16^ The *All of Us* Researcher Workbench is a secure cloud-based platform that allows approved researchers to use Jupyter Notebooks to access the data of participants that have consented to share their data within the Curated Data Repository (CDR).^16^

Inclusion criteria for the analytic sample were as follows: 1) Being 18+ years of age at the time of data collection, 2) Having an endometriosis diagnosis based on either electronic health record (EHR) data OR self-reported surveys (“*Has a doctor or health care provider ever told you that you have endometriosis?*”), which enable inclusion of a comprehensive cohort.^16^ This was also done because EHR data might not have been available for participants in the All of Us Research Program. 2) Having available Fitbit data on PA within the *All of Us* dataset. Upon enrollment, participants have the option to give permission to link their Fitbit devices with the Workbench, which allows use of their PA data from their trackers. The analytic sample therefore is limited to those for whom such data were available. While endometriosis is typically diagnosed during reproductive years, duration of its impact can vary across individuals. We therefore did not limit the upper age to allow extension into older age groups and enhance the age diversity in the sample.

### Study Variables

#### 1) Outcome Variable

We used daily step counts as the outcome variable based on several reasons. First, step counts capture all intensities and ambulatory types of PA, therefore providing the most comprehensive measure of habitual PA as per our *priori* goal. Second, there are published cut-points for step counts that have been linked to overall health outcomes, which allows their evaluation as a clinically meaningful, intuitive PA metric.^17,18,19^ Third, step counts are the most frequently used PA parameter in literature, which accordingly allows for easier comparison across studies.^20,21,22^

#### 2) Other PA Variables

To characterize the model-identified phenotypes by amounts of PA, we used light-, moderate-, vigorous-, and moderate-to-vigorous physical activity (MVPA) minutes recorded from wrist-worn Fitbit devices.^23,24^ MVPA was calculated by adding the moderate and vigorous intensity PA minutes.

#### 3) Demographic and disease-related factors

To describe the demographic characteristics of the study sample, we included age, ethnicity, race, employment, education level, and annual salary. In addition, to contextualize the identified cluster with respect to disease-relevant symptoms, we used the scores from the Patient-Reported Outcomes Measurement Information System (PROMIS) questionnaires^25,26^ on pain and fatigue symptoms. Pain and fatigue are among the two most common symptoms reported by individuals with endometriosis.^5^

The PROMIS Pain and Fatigue measures have been psychometrically evaluated in populations with chronic diseases.^25,26^ Pain is measured through the question “In the past 7 days, how would you rate your pain on average?” and the response options for PROMIS pain are numerical (Range= 0-10). For fatigue measurement, All of Us includes a single item from the PROMIS fatigue short form questionnaire (“In the past 7 days, how would you rate your fatigue on average?”). It provides categorical response options (i.e., “None”, “Mild”, “Moderate”, “Severe”, “Very Severe”, and “PMI: Skip”, indicating a skipped question within the AoU workbench.

### Statistical Analysis

The functional data analysis (FDA) framework for our study can be summarized in 4 main steps: 1) data organization and cleaning, 2) smoothing our data into functional form or data curves over a selected time period, 3) model selection, and 4) characterization of PA patterns and associations with related symptoms.

#### 1a) Data Organization and Cleaning

For the analysis of habitual PA patterns, we used 30 consecutive days of data (allowing up to 10 days of missingness) for each participant. All the 30 consecutive days per participant were selected within the months of spring (March, April, and May) to reduce the potential influence of seasonality on PA volume. To create the functional data matrix, each participant’s data was converted to wide format with 30 columns representing the 30 days of step count data, and 173 rows representing each participant’s data listed over the 30 days (173 x 30 data matrix).

#### 1b) Missing Data

The FMMs require all missing data to be imputed before model fitting. We imputed the days with missing PA data (up to 10 days per person within a 30-day span) using multiple imputation with chained equations (MICE) via predictive mean matching (PMM) from the *MICE* package in R.^27^ Multiple imputation is a well-established method for filling in missing data that has been applied in various fields and evaluated for its efficacy.^28,29,30^ We used 15 multiply imputed datasets and used the fraction of missing information (FMI) to evaluate the quality of the imputations.^31^ FMI provides a measure of the level of uncertainty of imputed data points in datasets with missingness^32^ and is a common method for evaluating decisions involving multiple imputation.^33^ We conducted steps 2 and 3 (outlined below) on all imputed datasets and pooled the results, as per previously established guidelines.^31,33^

#### 2) Data Smoothing (Conversion to Functional Data)

The participants’ imputed data matrices were smoothed using a Fourier-transform. This process involves decomposition of raw data into different frequency components to represent the time series data as a sum of sine and cosine functions. This connects our data points of daily step counts and forms a curve of step count activity over a 30-day period. Fourier transforms are considered a suitable application to data that vary over time and potentially have periodic behavior^34^ and demonstrated to perform well in previous similar studies.^12^

#### 3) Model Specification and Selection

For clustering, we implemented functional mixture models (FMMs) using the R *funFEM* library.^34^ FMMs constitute a probabilistic algorithm that predicts cluster membership to one of K clusters which are similar in nature. The model achieves this in a latent discriminative subspace where the ideal model results in the greatest discrimination *between* separate homogeneous clusters (K) while minimizing the variance between members *within* a cluster.^34^ The model assumes that the observed PA trajectories are generated from a mixture of underlying distributions, each representing a distinct cluster.^34^

Accordingly, FMMs were fit to the functional data matrix (described earlier) to identify latent clusters of PA trajectories. The optimal number of clusters was determined using the Bayesian information criterion (BIC). The BIC is a model selection criterion that balances a model’s fit with its complexity, penalizing overly complex models to prevent overfitting.^35^ We tested FMMs with up to 6 cluster (K) resolutions, and the model with the lowest BIC value was selected as the best-fitting model, representing the most appropriate number of clusters for the given data.

#### 4) Characterization of Model Phenotypes

After selecting the best-fitting model, we characterized the identified phenotypes `based` on the 30-day averages and their variability (SD) for all PA parameters. We used linear mixed regression models for pairwise comparisons where we regressed each of the PA parameters on the phenotypes, as well as participant as a random intercept. Next, we compared the PROMIS-based pain and fatigue scores across all clusters for a more disease-specific contextualization. We calculated the frequencies of responses for PROMIS pain and fatigue per cluster for our analysis. We used the *lmerTest* package in R for all regression analyses.^36^

## Results

### Sample Characteristics

Out of 825 eligible individuals with a diagnosis of endometriosis, 290 had Fitbit data available. The final analytic sample based on the allowable missingness in the given 30-day period included 173 participants, yielding 4,895 person-level days of data in total for analysis. The demographics of the study sample are provided in Table 1. The mean age was 59 years (standard deviation, SD = 14.8 years, Range = 23-90 years). Briefly, 153 (88.4%) self-identified as Not Hispanic or Latino, with greater than 81.5% (N = >141) self-identifying as White. Demographics were reported in this manner to comply with the All of Us Data and Statistics Dissemination Policy.^37^

**Table 1:**
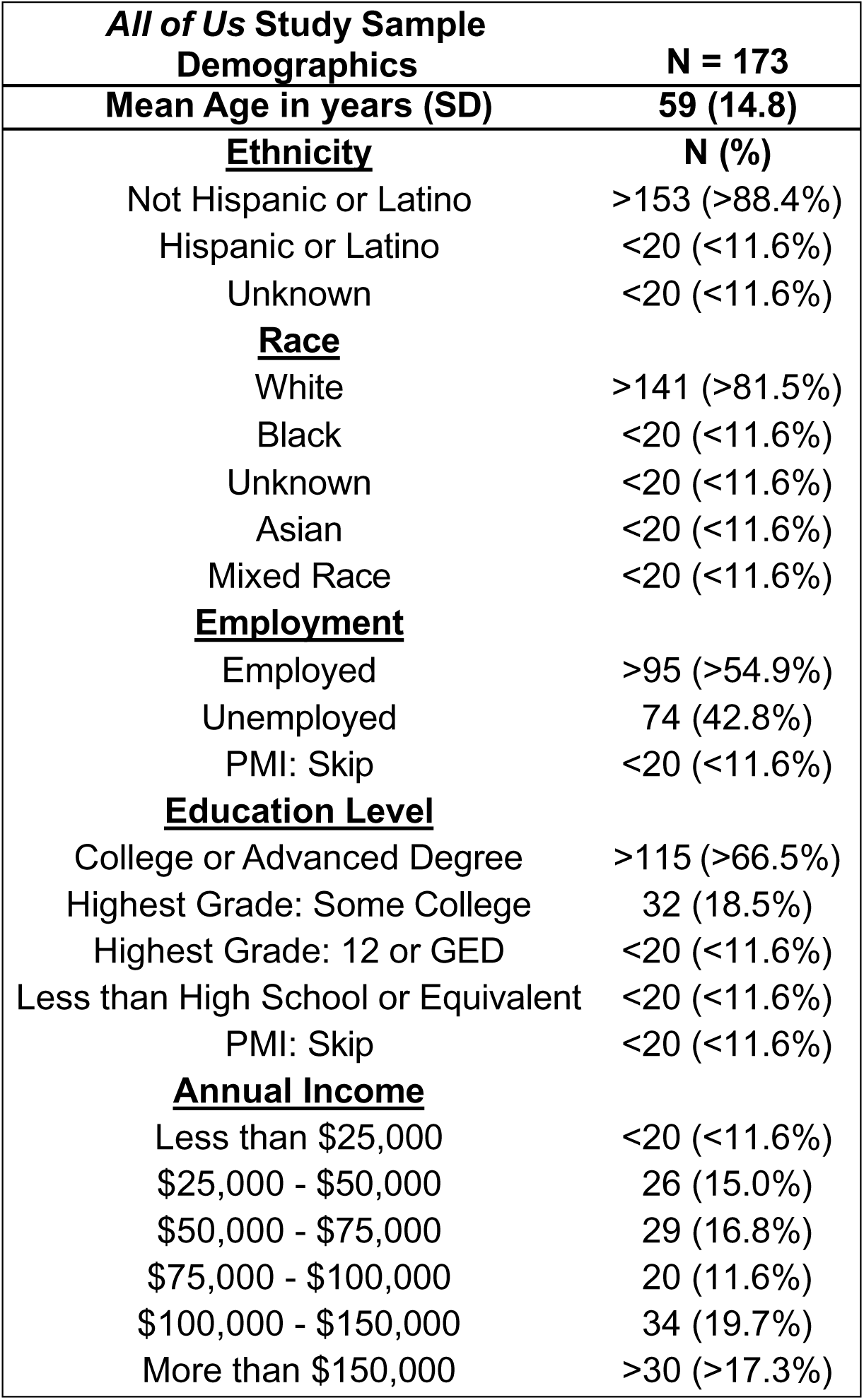
*All of Us* Study Sample Demographics. Overview of cohort demographics including mean age (SD), ethnicity, race, employment, education level, and annual income (N(%)). “<20” is used to remain in compliance with the *All of Us Research Program* Data Dissemination Policy, which states that N’s of <20 cannot be reported in tables or figures. “PMI: Skip” indicates that the participant skipped this question.

### Model Results

The model that provided the best fit for the data was a 4-cluster resolution (K=4) using a Fourier basis (Mean Δ BIC = 15.02). These clusters represent different patterns of PA (i.e., step counts) among women with endometriosis, with differences in PA volumes and variability over time. Supplemental Figure 1 depicts each participant’s orientation within the 4 clusters based on the model coefficients in the discriminative space of the FMM. Figures 1 and 2 respectively show the person– and cluster-level Fourier-smoothed curves of step count over 30 days.

**Figure 1:**
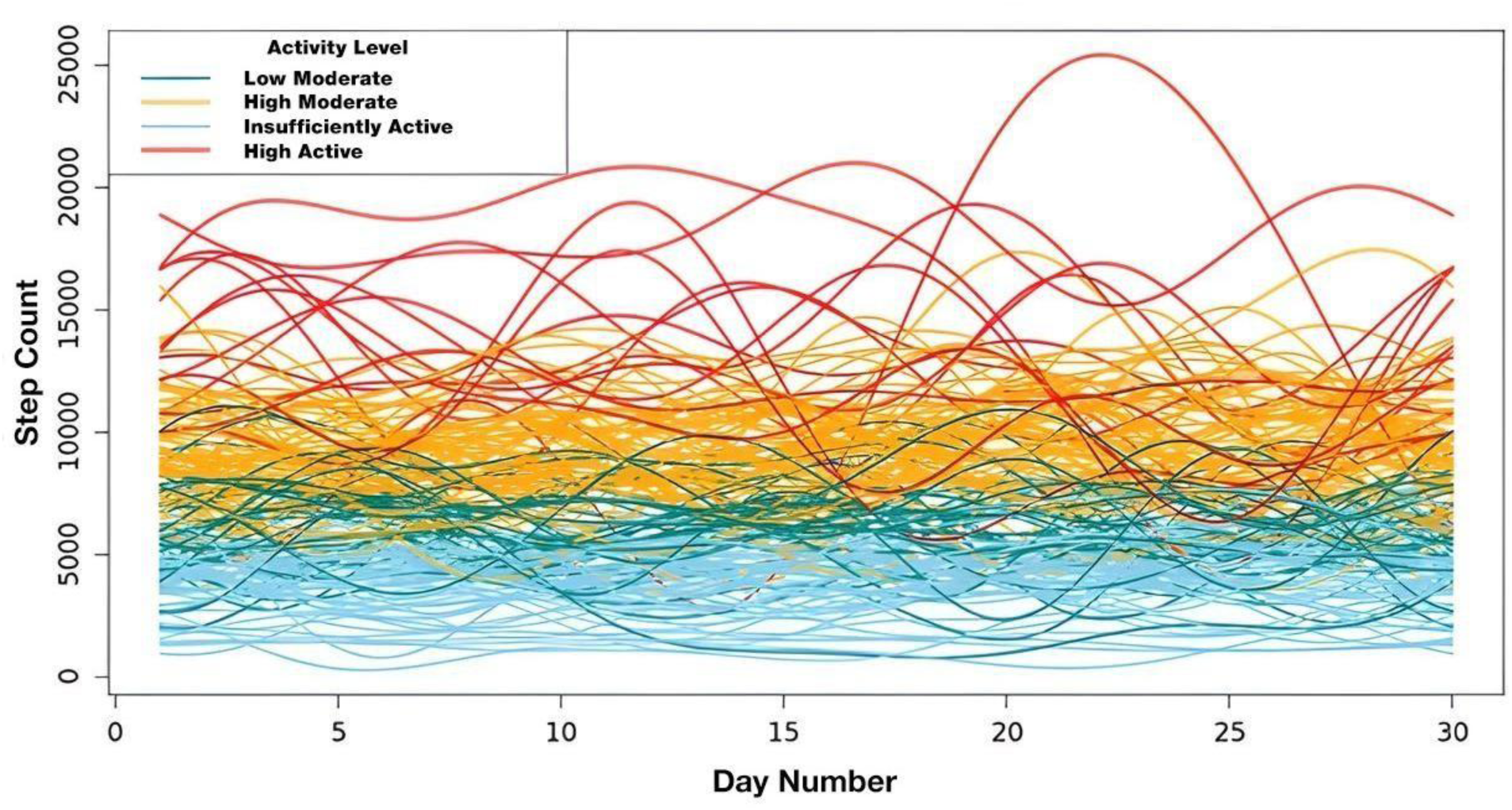
Smoothed Curves of Daily Step Count Data by Cluster. Fourier basis functions representing the daily step count activity of each participant (N=173) smoothed over 30 days. Red curves indicate the “High Active” cluster, orange curves indicate the “High Moderate” cluster, green curves indicate the “Low Moderate” cluster, and light blue curves indicate the “Insufficiently Active” cluster.

**Figure 2:**
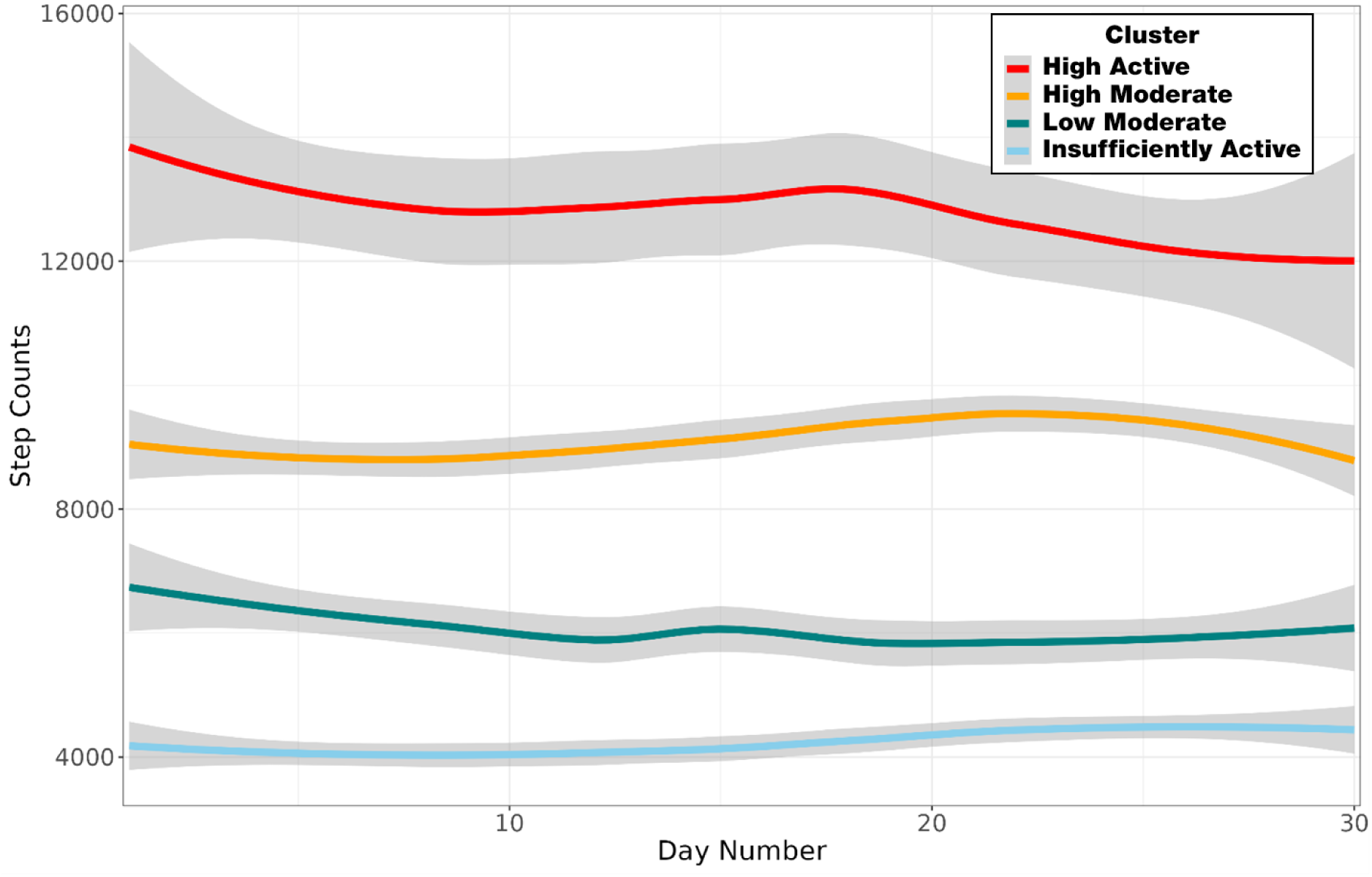
Average Smoothed Curves of Daily Step Counts by Cluster. Mean of the smoothed curves of step counts for the functional mixture model (FMM)-identified clusters over the 30-day period. Red curve indicates the “High Active” cluster, orange curve indicates the “High Moderate” cluster, green curve indicates the “Low Moderate” cluster, and light blue curve indicates the “Insufficiently Active” cluster. Gray areas indicate 95% confidence intervals.

### Cluster Characterization

Cluster-level descriptive statistics on all PA parameters are provided in Table 2. Based on our analysis, we refer to these 4 clusters as *‘High Active’, ‘High Moderate’, ‘Low Moderate’ and ‘Insufficiently Active”*. The first cluster – “*High Active*” – comprised the smallest cluster (*n*=16) and was associated with the highest daily mean steps and PA minutes for all intensities. Of note, this cluster was also associated with the highest day-to-day variance in steps. The second cluster *-“High Moderate”* – was the largest cluster (*n*=64) and had fewer daily steps compared to “*High Active*” but similar minutes of MVPA (i.e., 60.1 vs 70.8 minutes, respectively, *p*=0.292). The third cluster – “*Low Moderate*” (*n*=25) – had lower daily steps and minutes of PA in comparison to *High Moderate*, but similar day-to-day variance (See Figure 2 and 5). This cluster overall had slightly higher volumes for all PA intensities in comparison to the fourth cluster, along with higher fluctuations (See table 2). The fourth cluster – “*Insufficiently Active*” – was the second largest (*n*=58) and was characterized by the least number of daily steps and minutes of all PA intensities. Of note, this cluster had the least amount of day-to-day variance for all PA parameters. We provide boxplots depicting the distributions of day-level step counts and all intensities of PA for these clusters in Supplemental Figure 2A-E. Comparisons of daily lightly active and MVPA minutes are shown in Figure 3 where the overall trend of reduced volume and variability is visible as the clusters move from most active (“*High* Active”) to least active (“*Insufficiently Active*”).

**Figure 3:**
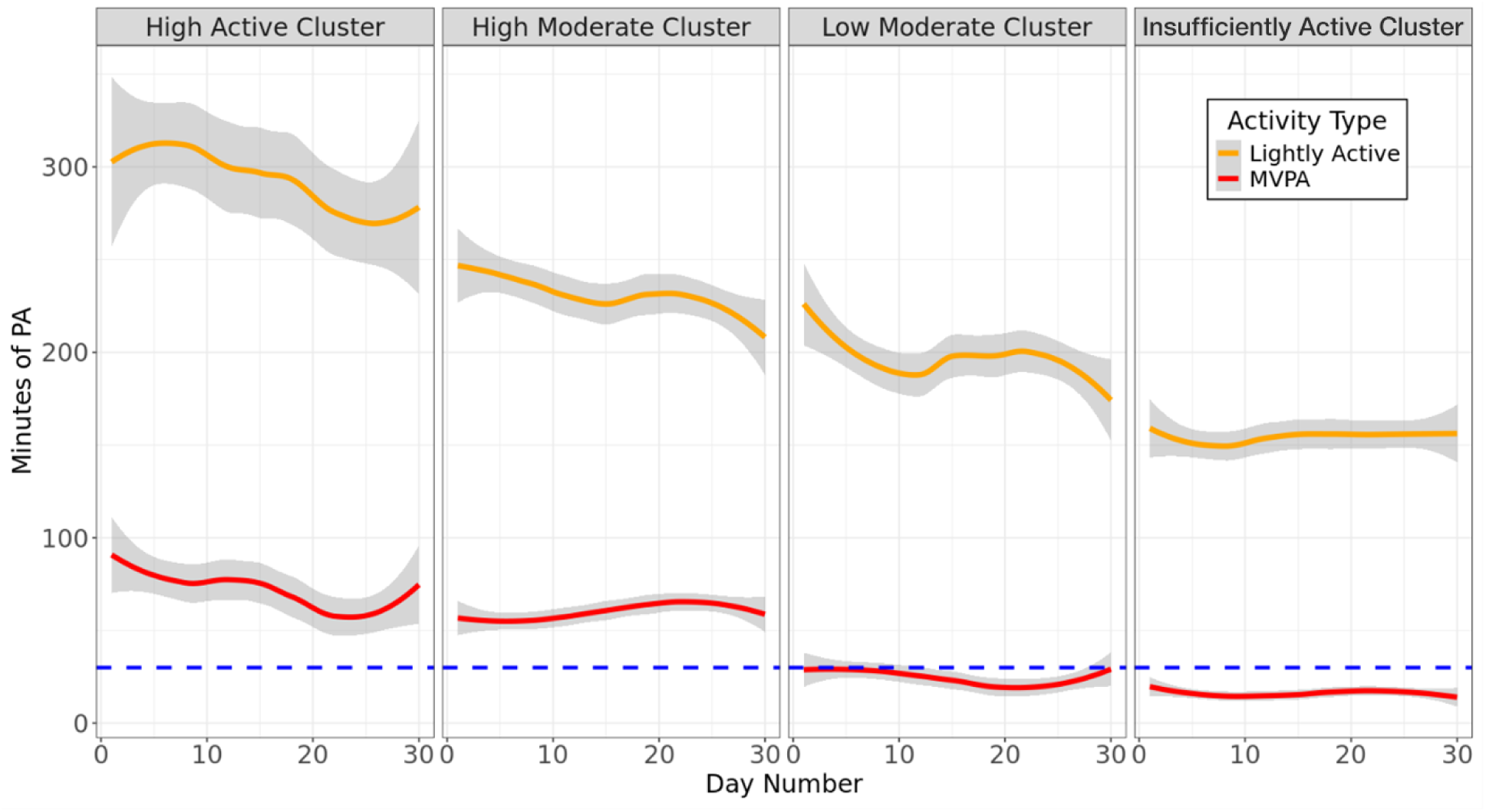
Daily Lightly Active and MVPA Patterns by Cluster. Cluster-level means of daily lightly active minutes and moderate-to-vigorous (MVPA) minutes over 30 days. Red curves indicate MVPA minutes. Orange curves indicate lightly active minutes. Grey areas surrounding curves indicate 95% confidence intervals around lightly active and MVPA minutes over the 30-day period. Blue dashed line indicates the daily recommended MVPA amount of 30 minutes.

**Table 2:** Cluster-level Daily Step Counts and Physical Activity Minutes Means and Ranges. Cluster-level daily step count and activity minutes means (Cluster 4 “*Insufficiently Active*”: N = 58, Cluster 3 “*Low Moderate*”: N = 25, Cluster 2 “*High Moderate*”: N = 64, Cluster 1 “*High Active*”: N = 16). Bold in table indicates physical activity variable for each cluster comparison.

| Physical Activity Variable | Day-Level Mean (SD) | Range |
| --- | --- | --- |
| **Step Counts** |  |  |
| Cluster 4 (Insufficiently Active) | 4235.7 (2174.7) | 102 – 14349 |
| Cluster 3 (Low Moderate) | 6067.5 (2797.0) | 119 – 17693 |
| Cluster 1 (High Moderate) | 9125.4 (3631.0) | 106 – 26486 |
| Cluster 2 (High Active) | 12777.0 (5248.2) | 390 - 29368 |
| **Lightly Active Minutes** |  |  |
| Cluster 4 (Insufficiently Active) | 154.0 (87.6) | 0-468 |
| Cluster 3 (Low Moderate) | 196.7 (88.5) | 0-726 |
| Cluster 1 (High Moderate) | 231.5 (128.9) | 0-1440 |
| Cluster 2 (High Active) | 293.0 (142.6) | 0-796 |
| **Fairly Active Minutes** |  |  |
| Cluster 4 (Insufficiently Active) | 11.3 (23.2) | 0 – 242 |
| Cluster 3 (Low Moderate) | 15.2 (25.1) | 0 – 191 |
| Cluster 1 (High Moderate) | 40.4 (50.3) | 0 – 335 |
| Cluster 2 (High Active) | 43.9 (48.7) | 0 – 234 |
| **Very Active Minutes** |  |  |
| Cluster 4 (Insufficiently Active) | 4.8 (10.3) | 0 – 114 |
| Cluster 3 (Low Moderate) | 9.1 (18.4) | 0 – 164 |
| Cluster 1 (High Moderate) | 19.7 (25.5) | 0 – 192 |
| Cluster 2 (High Active) | 26.9 (33.1) | 0 - 160 |
| **MVPA Minutes** |  |  |
| Cluster 4 (Insufficiently Active) | 16.1 (25.4) | 0 – 356 |
| Cluster 3 (Low Moderate) | 24.3 (31.1) | 0 – 355 |
| Cluster 1 (High Moderate) | 60.1 (56.4) | 0 – 527 |
| Cluster 2 (High Active) | 70.8 (58.9) | 0 - 394 |

### Characterization of model identified phenotypes

Results from the mixed-level regression models for post-hoc comparison among the identified clusters on different PA intensities are provided in Table 3. All point estimates for step counts and light intensity PA minutes were statistically significant (*p*<0.05; See Table 3). For MVPA minutes, the *Low Moderate* and *Insufficiently Active* phenotypes were associated with significantly fewer minutes than *High Moderate* (*p*<0.05 for both), but the difference in MVPA between *High Active* and *High Moderate* was not statistically discernable (*p*= 0.29).

**Table 3:** Linear Mixed Regression Model Results Comparing Step Counts, Lightly Active and MVPA Minutes Between Clusters. Results of linear mixed regression models comparing mean daily step count, mean daily lightly active minutes, and mean daily moderate-to-vigorous (MVPA) across clusters. Comparisons are between Clusters 2 and 1 (“High Moderate” and “High Active”), Clusters 2 and 3 (“High Moderate” and “Low Moderate”), and Clusters 2 and 4 (“High Moderate” and “Insufficiently Active”). All adjusted p-values for comparisons are significant (p<0.05) indicated by the “*”, except for the comparison of MVPA minutes between “High Moderate” and “High Active”.

| PA Variable | Comparison Groups | Estimate | Error | t-statistic | df | Adjusted p-value |
| --- | --- | --- | --- | --- | --- | --- |
| Step Counts | Cluster 2 (Intercept) | 9,123.7 | 153.3 | 59.53 | 167.6 | <0.001 |
|  | Clusters 2-1 | +3,678.4 | 343.6 | 10.71 | 169.2 | <0.001 |
|  | Clusters 2-3 | -3,091.0 | 280.9 | -11.00 | 169.1 | <0.001 |
|  | Clusters 2-4 | -4,902.2 | 225.6 | -21.73 | 169.8 | <0.001 |
| Lightly Active Minutes | Cluster 2 (Intercept) | 231.2 | 9.7 | 23.90 | 168.9 | <0.001 |
|  | Clusters 2-1 | +61.6 | 21.6 | 2.84 | 169.2 | 0.005 |
|  | Clusters 2-3 | -36.8 | 17.7 | -2.08 | 169.2 | 0.0392 |
|  | Clusters 2-4 | -78.5 | 14.2 | -5.52 | 169.3 | <0.001 |
| MVPA Minutes | Cluster 2 (Intercept) | 60.1 | 4.5 | 13.34 | 169.1 | <0.001 |
|  | Clusters 2-1 | +10.7 | 10.1 | 1.06 | 169.3 | 0.292* |
|  | Clusters 2-3 | -36.1 | 8.2 | -4.38 | 169.3 | <0.001 |
|  | Clusters 2-4 | -44.3 | 6.6 | -6.69 | 169.4 | <0.001 |

### Comparisons of reported pain and fatigue levels by cluster

Figures 4 and 5 depict the cluster-level scores from the PROMIS pain and fatigue questionnaires, respectively. The PROMIS pain scores by cluster are depicted in boxplots in Figure 4 and the mean pain scores were as follows: *Active*: 1.81, *High Moderate*: 2.33, *Low Moderate*: 3.48, and *Insufficiently Active*: 3.69. The proportions of fatigue responses within a cluster are provided in Figure 5. Both the *Insufficiently Active* and *High Active* clusters had relatively higher proportions of “Severe” fatigue (i.e., 11.9% and 11.8%, respectively) compared to other phenotypes. The *High Active* phenotype was further associated with highest proportion of “None” for fatigue levels (23.5%). The *Low Moderate* and *High Moderate* phenotypes had the highest combined proportions of “None” or “Mild” fatigue levels at 63.2% and 62.1%, respectively (see Figure 5).

**Figure 4:**
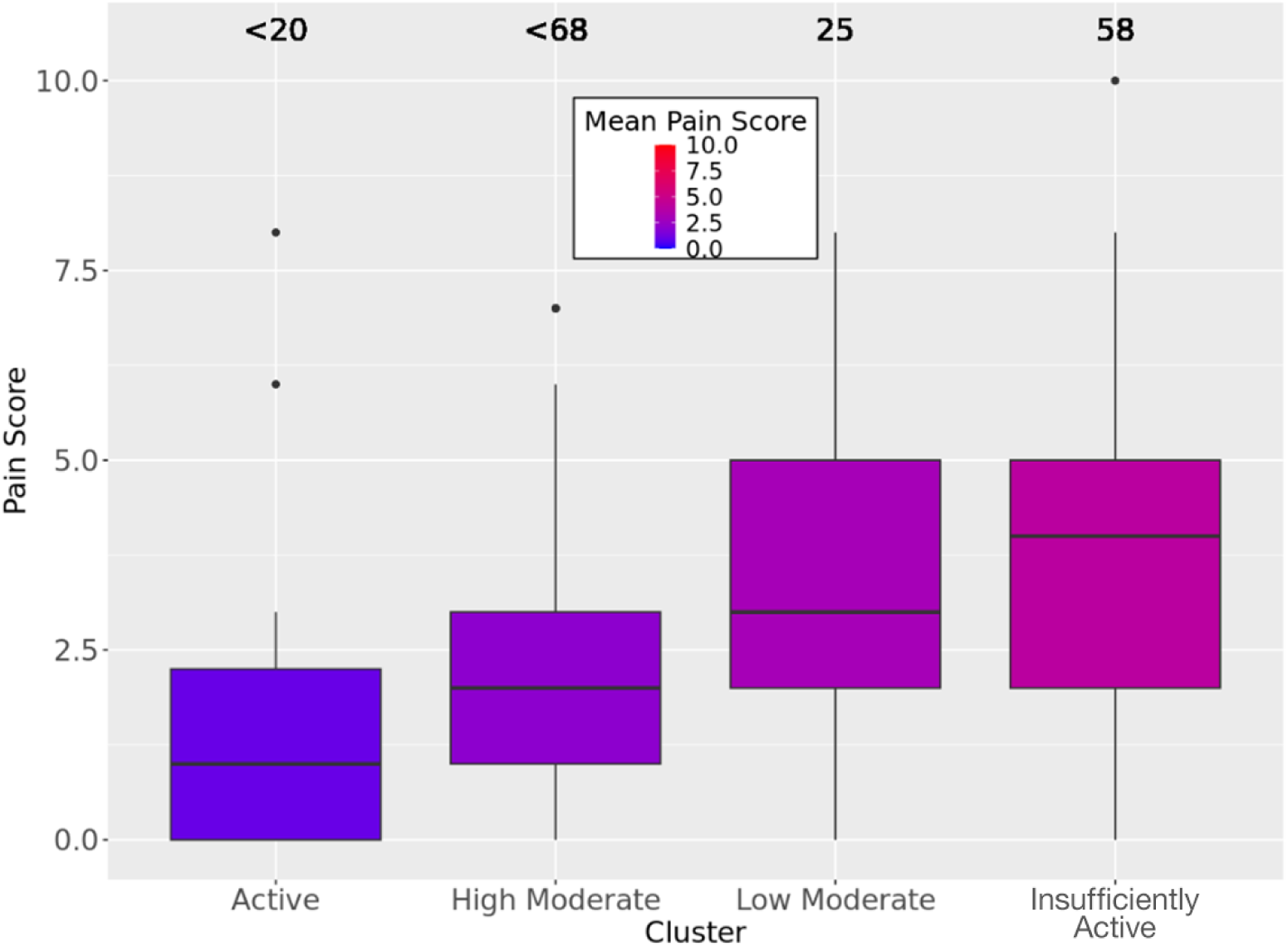
PROMIS Pain Score Distribution by Cluster. Mean PROMIS pain scores by cluster. Pain scores range from 0-10 as indicated in the legend (using a scale from blue (0) to red (10)). Boxplot colors correspond to color gradient of pain score responses. N displayed above each boxplot. “<20” is used to remain in compliance with the *All of Us Research Program* Data Dissemination Policy, which states that N’s of <20 cannot be reported in tables or figures. Boxplot shows the range of PROMIS pain scores in each cluster with the solid black line through the boxplots indicating the mean PROMIS pain score.

**Figure 5:**
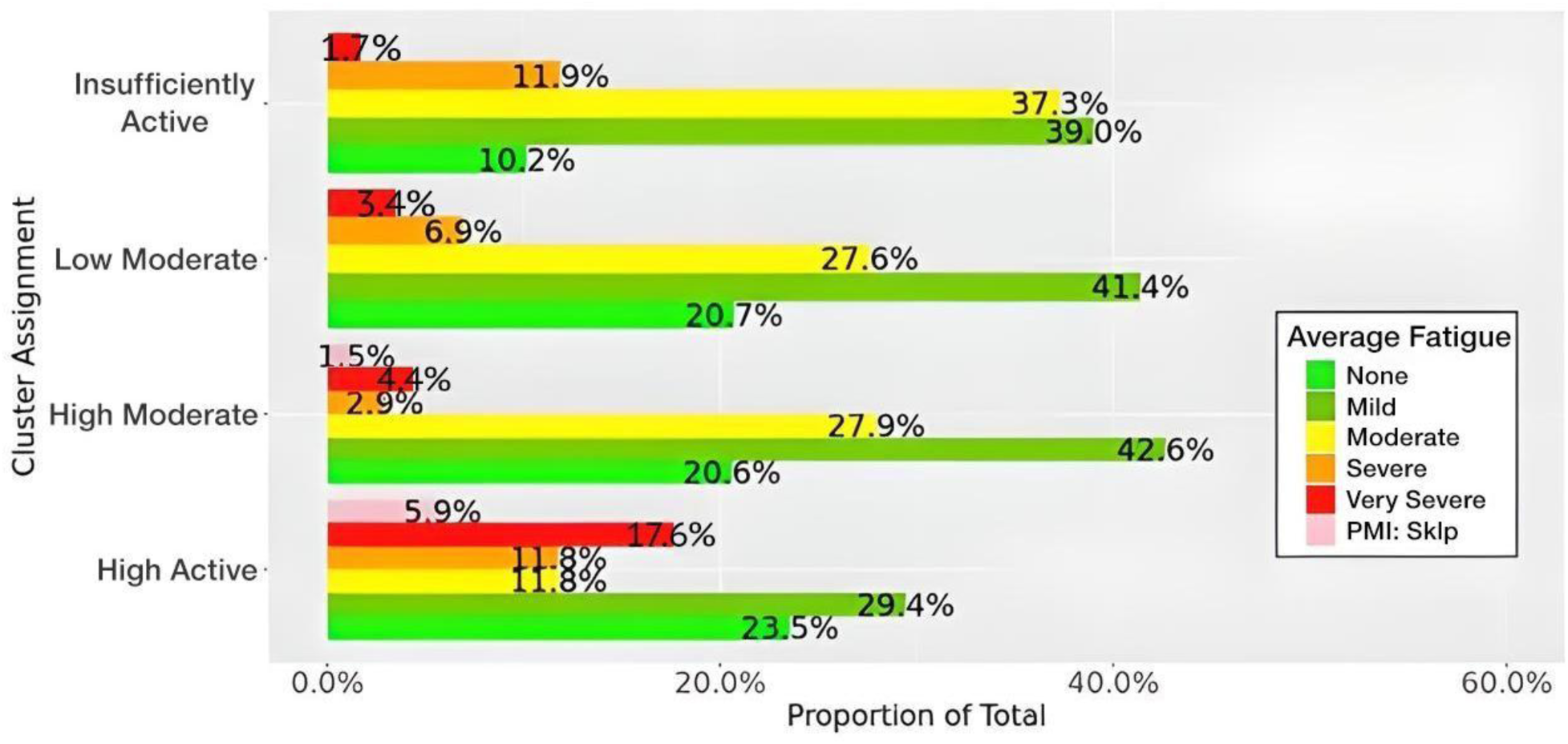
Proportion of PROMIS Fatigue Responses by Cluster. PROMIS fatigue responses by cluster as a proportion of the total number of participants in each cluster assignment. “PMI: Skip” indicates that the participant skipped the question, or no response was collected for PROMIS fatigue.

## Discussion

In this study, we investigated PA trajectories over a 30-day period in a sample of women with endometriosis. We used the FDA framework to investigate the PA trajectory clusters, which leverages data curves instead of aggregated data points. This retains more information (e.g. temporal trends) than otherwise, enabling a more comprehensive analysis of PA patterns. Our results indicated 4 distinct phenotypes characterized by differences in PA volume and variability. Further, self-reported pain and fatigue levels varied among the phenotypes, pointing to possible beneficial effects of PA on pain. Our findings highlight the potential of analyzing PA trajectories using a functional data framework, especially in a population impacted by chronic pain.

The results from the best fitting model indicated 4 distinct phenotypes of PA patterns based on daily step counts, further characterized by differences in total daily minutes and intra-day variances in the PA parameters. The *High Active* cluster was characterized by the highest volume of PA and had the highest variability over the 30-day period. The *High Active* phenotype was associated with the highest average MVPA minutes among all phenotypes with 70.8 minutes a day. The *High Moderate* phenotype was characterized by a relatively lower average step count (See Table 2), but similar daily MVPA minutes compared to the *High Active* phenotype. The *Low Moderate* phenotype was characterized by a lower average step count than the *High Moderate* cluster, but a substantial decrease in average MVPA minutes over time. The *Insufficiently Active* phenotype was characterized by the lowest average step count (4235.7 steps/day) and lowest average MVPA minutes. The *Insufficiently Active* phenotype was also associated with the lowest variance for step count over the 30-day period. Our step count findings are consistent with previous research on special populations (such as older individuals and those with chronic illnesses), which report daily step counts ranging from 1,200 to 8,800 steps.^38^ Our analyses suggest that the *Insufficiently Active* and *Low Moderate* phenotypes are more likely to fall within this range, while the *High Moderate* and *High Active* phenotypes are more likely to exceed it.

Although the previous study on special populations did not specifically include women with endometriosis, our results suggest that some individuals in this population surpass the expected PA levels for those with chronic illnesses (i.e. diabetes, breast cancer, fibromyalgia, arthritis, neuromuscular diseases).

In the *High Active* phenotype, the average daily MVPA minutes (i.e., 70.8 minutes) was twice the daily recommended amount (30 min/day) by the U.S. Department of Health and Human Services (HHS).^39^ The *High Moderate* phenotype was similarly associated with 60.1 mins/day of MVPA. These findings suggest that women in the *High Active* and *High Moderate* phenotypes are most likely to accumulate at least the minimum amount of PA recommended for overall health on most days. In contrast, the *Low Moderate* and *Insufficiently Active* phenotypes were associated with significantly lower daily MVPA mins (i.e., 24.3 and 16.1 mins/day, respectively). While only the *Insufficiently Active* phenotype does not meet the weekly threshold of 150 min/week set by the HHS, the *Low Moderate* phenotype does meet this threshold at 170.1 min/week. The PA patterns of these phenotypes also indicated lower variability in PA over time, suggesting these individuals tend to remain consistent within their respective lower activity patterns.

Our analyses further indicated that PROMIS pain scores were inversely related to PA volume where *High Active* phenotype was associated with the lowest pain levels based on the PROMIS questionnaire. A cross-sectional cohort study comparing PROMIS 7-day pain intensity between controls and chronic conditions (osteoarthritis, premenstrual syndrome, hernia repair, and breast cancer) demonstrated that mean pain intensity for the control group was ∼2 on the 0-10 scale.^40^ In comparison, the individuals in our *High Active* phenotype have lower scores (i.e., 1.7), the *High Moderate* phenotype has similar levels of pain to controls, whereas the scores for the *Low Moderate* and *Insufficiently Active* phenotypes are higher. This provides further support to the notion that increased habitual PA may play a role in mitigating pain associated with endometriosis. Our findings are further in line with a review supporting the underlying physiological mechanisms for PA related analgesia, which lends biological plausibility to our results.^41^ This is noteworthy given that previous research indicates a strong association between regular MVPA and improved symptom management and quality of life in individuals with chronic conditions, including endometriosis.^5,7,42,43,44^ Accordingly, our findings provide further support to the notion that increased habitual PA may play a role in mitigating pain associated with endometriosis. To our knowledge, this is the first study reporting this association in endometriosis based on trends of PA using objectively estimated data.

In contrast to pain scores, the trends in PROMIS fatigue scores indicated variable associations with the phenotypes. For example, while the *High Active* phenotype was associated with the most “None” responses amongst the 4 phenotypes, it also had the same proportion of “Very Severe” responses as the *Insufficiently Active* phenotype (i.e., ∼11%; See Figure 7). On the other hand, *Low Active* and *Moderately Active* phenotypes were associated with the highest frequencies of “Mild” responses.

The *Insufficiently Active* phenotype additionally had the highest prevalence of “Moderate” fatigue responses. The *Insufficiently Active* phenotype had the lowest prevalence of no fatigue, while the *High* Moderate phenotype was associated with the lowest prevalence of “Severe” fatigue. In sum, these trends point to a beneficial effect, but fatigue can be difficult to measure and capture^45^ therefore more research needs to be done to confirm this interaction.

Although research on clustering PA behavior among chronic disease populations is limited, our study shares some similarities with those from O’Regan et al.^46^, who also identified four PA patterns using accelerometry-based step counts in participants with high cholesterol, hypertension, arthritis, and other circulatory conditions: “inactive-sedentary,” “low activity,” “active,” and “very active.” Their average step counts were 4,328, 8,158, 12,681, and 17,982 steps/day, respectively, with MVPA averages of 7.7, 25.5, 52.2, and 94.4 minutes/day. Our findings, which also identified four clusters, indicate that the *Insufficiently Active* and *Low Moderate* phenotypes averaged fewer than 5,000 steps/day, with the two lowest activity clusters not meeting the 30 minutes/day HHS recommendation for MVPA. In line with O’Regan et al., 2021^46^, who reported that lower PA levels were associated with higher morbidity and healthcare utilization, our results support the notion that women with lower PA levels are more likely to face greater health challenges, consistent with previous literature on chronic diseases.

These results collectively point to the complexity of the relationship between PA and disease symptoms, such as pain and fatigue. While the *High Active* phenotype had the greatest levels of PA, they recorded a substantial proportion of “Very Severe” and “Severe” fatigue even though their pain levels were the lowest among all phenotypes. This varied relationship between PA and pain and fatigue potentially highlights that increased PA can reduce pain but does not necessarily decrease fatigue levels in this population. Alternatively, optimal ranges of PA could vary based on individual needs to maximize its benefits for endometriosis symptom management (i.e., reduce or prevent pain without exacerbating fatigue). It is possible that for some individuals, a consistent distribution of PA, as opposed to intense bouts of PA, is more efficacious for flare-up prevention.^6^ Similarly, regularity might be more important than the intensity in this context.^5^ Future studies should further investigate this relationship in larger samples, with additional disease-specific and temporally aligned self-report data to further contextualize PA behavior.

Finally, the observed age distribution (spanning several decades) presents a unique opportunity to study PA maintenance across different life stages among those with endometriosis. Given the paucity of research across the lifespan among those with endometriosis, the age diversity in this study aligns with the All of Us cohort’s intentional inclusion of underrepresented older populations in chronic disease research. It also strengthens generalizability to clinical populations while highlighting the need for age-specific PA recommendations in chronic gynecological conditions. Nevertheless, more focused future studies are warranted to stratify analyses by menopausal status, as estrogen fluctuations may differentially impact both endometriosis symptoms and PA patterns.

## Limitations

This study has several limitations. First, we use retrospective, secondary data from the *All of Us Research Program*, which uses self-report surveys and these do not include some of the variables that could potentially be relevant to our analysis, such as habitual PA behaviors, other health behaviors, menstrual cycle or health, or pain medication use at the time that corresponds to the Fitbit-based PA data. As such, our ability to contextualize the phenotypes was limited to the available variables (e.g., pain, fatigue) for assessing symptom fluctuations and their relationship to PA. Second, we used 30 consecutive days of data limited to spring months (March, April, May) to reduce potential seasonal influence and introduce some level of standardization across individuals. This was also based on data availability, i.e., these months had more data and less intermittent missingness overall compared to other months. However, we acknowledge this hinders us from examining seasonal variations in PA patterns that could be relevant. Third, ∼82% of participants identified as White and ∼88% as non-Hispanic, restricting the generalizability of findings to more diverse populations.

Similarly, our analysis is limited to individuals who own Fitbit devices and consented to data sharing, introducing potential bias where the sample may represent individuals who are already more active. This would suggest a possible overestimation of the PA levels in our population of interest. Accordingly, future studies can focus on prospective PA data collection striving to enroll participants from diverse backgrounds. This would enable more comprehensive identification of those who stand to benefit the most from targeted, personalized PA for symptom management, especially in understudied populations (minority races/ethnicities and sexual and gender minorities).

## Conclusion

This study demonstrates the utility of functional data analysis (FDA) in characterizing PA trajectories among women with endometriosis, revealing insights into the complex relationship between PA volume, intensity, and symptom management in a chronic pelvic pain population. The results from the FMMs based on step counts revealed 4 distinct phenotypes of PA patterns. These phenotypes displayed different magnitudes of variance over the 30-day period in PA parameters, in addition to total volume. There was also a significant association between decreased likelihood of pain with increasing volumes of PA, suggesting its protective potential against disease-related pain. Phenotyping enables identification of those at greater risk for physical inactivity and who could benefit most from personalized intervention approaches to improve PA behavior. A nuanced approach to personalized PA recommendations could improve quality of life in this population and more studies are needed to investigate this relationship. These insights collectively underscore the potential benefits of promoting personalized PA interventions, particularly for women aiming to leverage PA as part of their endometriosis management plan.

## Data Use and Availability

Data used for this manuscript are obtained from the *All of Us Research Program* and not owned by the authors. Data from the *All of Us Research Program* are accessible only through the Researcher Workbench (https://workbench.researchallofus.org), as stipulated in the informed consent of participants in the program. This data use agreement prohibits investigators from providing row level data on *All of Us* participants and thus providing a de-identified dataset is not possible for this manuscript. The *All of Us Data and Statistics Dissemination Policy* also stipulates that “No data or statistics can be reported that allow a participant count of 1 to 20 to be derived from other reported cells or information, including in text, tables, or figures. This includes the use of percentages or other mathematical formulas that in combination would allow an individual to deduce a participant count of less than 20” requiring participant counts less than 20 to be reported as <20 in tables and figures. This study was completed in compliance with all of the policies outlined in the “Overview of All of Us Research Program Policies for Researchers” (https://support.researchallofus.org/hc/en-us/articles/4415498292244-Overview-of-All-of-Us-Research-Program-Policies-for-Researchers). The code used for this demonstration project is available within the Researcher Workbench at https://workbench.researchallofus.org/workspaces/aou-rw-98708e1d/femalereproductivedisordersacrossdiversepatientpopulations/data. Any investigator interested in accessing data used for this manuscript or any other All of Us Research Program data can do so following the procedures outlined in https://www.researchallofus.org/apply/.

## Data Availability

Data used for this manuscript are obtained from the All of Us Research Program and not owned by the authors. Data from the All of Us Research Program are accessible only through the Researcher Workbench (https://workbench.researchallofus.org), as stipulated in the informed consent of participants in the program. This data use agreement prohibits investigators from providing row level data on All of Us participants and thus providing a de-identified dataset is not possible for this manuscript. The All of Us Data and Statistics Dissemination Policy also stipulates that no data or statistics can be reported that allow a participant count of 1 to 20 to be derived from other reported cells or information, including in text, tables, or figures. This includes the use of percentages or other mathematical formulas that in combination would allow an individual to deduce a participant count of less than 20, requiring participant counts less than 20 to be reported as less than 20 in tables and figures. This study was completed in compliance with all of the policies outlined in the Overview of All of Us Research Program Policies for Researchers (https://support.researchallofus.org/hc/en-us/articles/4415498292244-Overview-of-All-of-Us-Research-Program-Policies-for-Researchers). The code used for this demonstration project is available within the Researcher Workbench at https://workbench.researchallofus.org/workspaces/aou-rw-98708e1d/femalereproductivedisordersacrossdiversepatientpopulations/data. Any investigator interested in accessing data used for this manuscript or any other All of Us Research Program data can do so following the procedures outlined in https://www.researchallofus.org/apply/.

## Acknowledgements

The All of Us Research Program is supported by the National Institutes of Health, Office of the Director: Regional Medical Centers: 1 OT2 OD026549; 1 OT2 OD026554; 1 OT2 OD026557; 1 OT2 OD026556; 1 OT2 OD026550; 1 OT2 OD 026552; 1 OT2 OD026553; 1 OT2 OD026548; 1 OT2 OD026551; 1 OT2 OD026555; IAA #: AOD 16037; Federally Qualified Health Centers: HHSN 263201600085U; Data and Research Center: 5 U2C OD023196; Biobank: 1 U24 OD023121; The Participant Center: U24 OD023176; Participant Technology Systems Center: 1 U24 OD023163; Communications and Engagement: 3 OT2 OD023205; 3 OT2 OD023206; and Community Partners: 1 OT2 OD025277; 3 OT2 OD025315; 1 OT2 OD025337; 1 OT2 OD025276. We gratefully acknowledge All of Us participants for their contributions, without whom this research would not have been possible. We also thank the National Institutes of Health’s All of Us Research Program for making available the participant data [and/or samples and/or cohort] examined in this study.

