## Supplemental Materials for "Physical Activity Phenotypes in Endometriosis Using Unsupervised Learning via Functional Mixture Models"

**Supplemental Figure 1: FMM Coefficients in Discriminative Space Within Model**

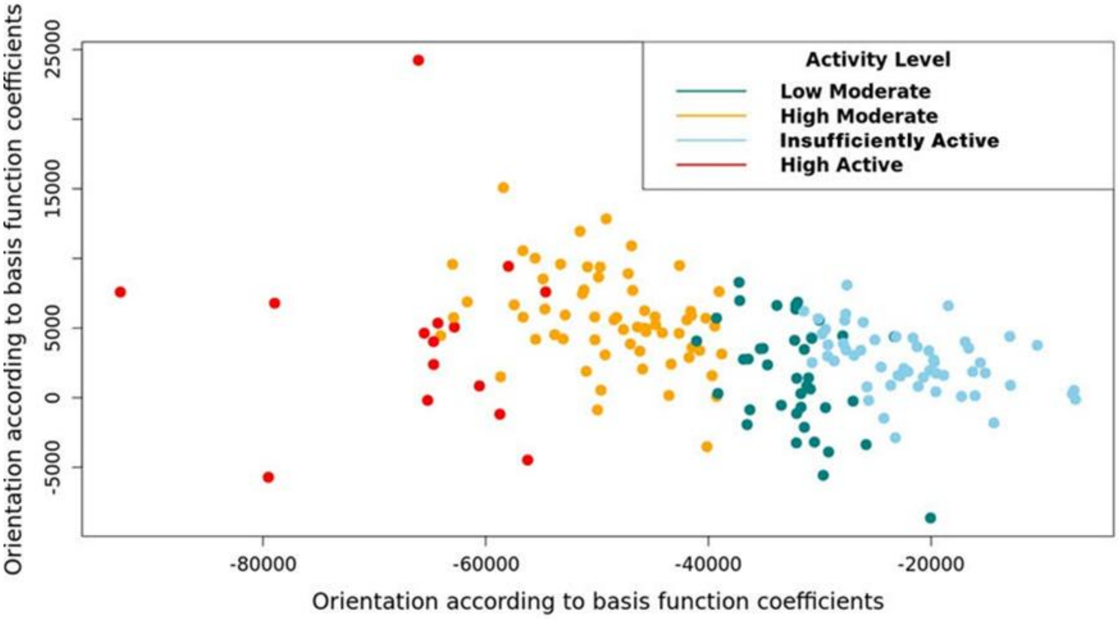


**Supplemental Figure 1:** Individual functional mixture model (FMM) coefficients within the discriminative space of the clustering model. Each dot represents 1 participant (N=173). Red dots indicate the "High Active" cluster, orange dots indicate the "High Moderate" cluster, green dots indicate the "Low Moderate" cluster, and light blue dots indicate the "Insufficiently Active" cluster.

**Supplemental Fig. 2A: Day-Level Average Step Counts**

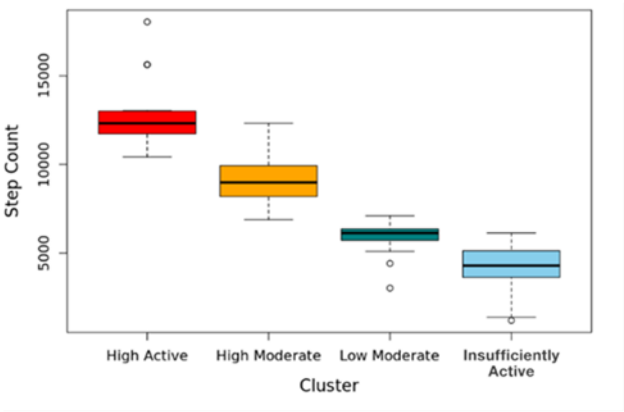


**Supplemental Fig. 2B: Day-Level Average Light Intensity PA**

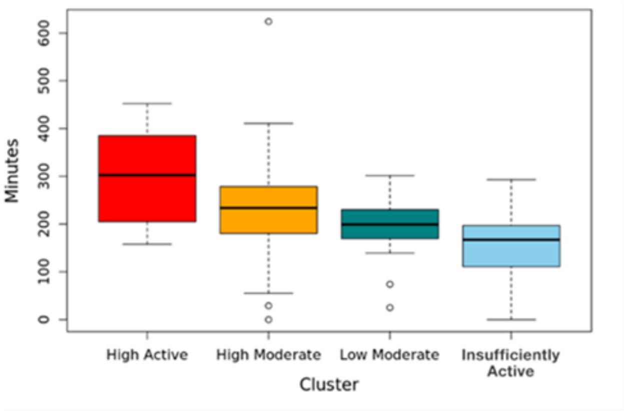


**Supplemental Fig. 2C: Day-Level Average Moderate Intensity PA**

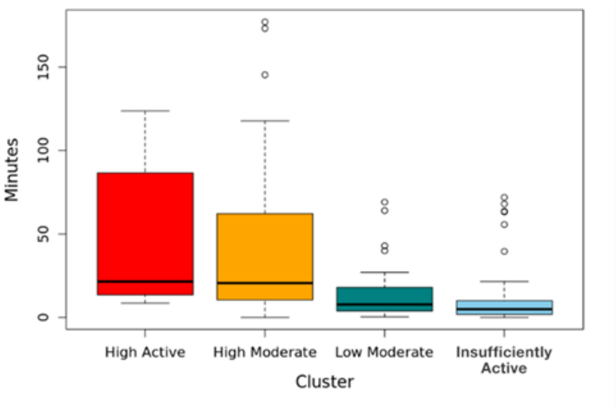


**Supplemental Fig. 2D: Day-Level Average MVPA**

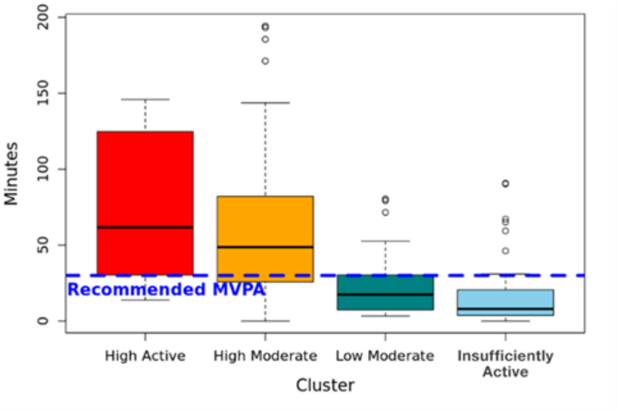


**Supplemental Fig. 2E: Day-Level Average Vigorous Intensity PA**

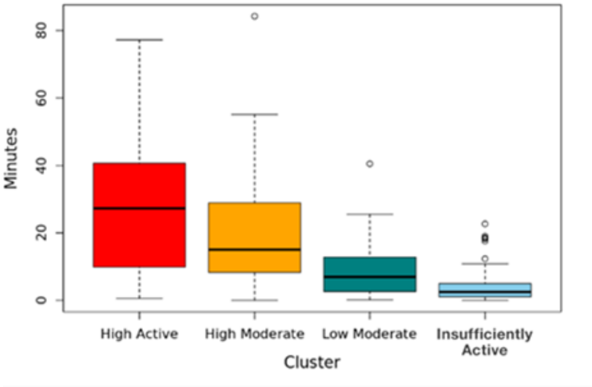


**Supplemental Figure 2 (A-E):** Boxplots of day-level step counts (4A) and physical activity (PA) minutes of all intensities (light, moderate, moderate-to-vigorous (MVPA), and vigorous – 4B-4E) across all clusters. Red boxplot indicates the "High Active" cluster, orange boxplot indicates the "High Moderate" cluster, green boxplot indicates the "Low Moderate" cluster, and light blue boxplot indicates the "Insufficiently Active" cluster. Solid black line through boxplots show means for each parameter. Error bars included with each boxplot.

**Supplemental Table 1: Fraction of Missing Information after Multiple Imputation of Daily Step Count Datasets**

| Variable | FMI |
|---|---|
| Day 1 | 0.076 |
| Day 2 | 0.091 |
| Day 3 | 0.075 |
| Day 4 | 0.060 |
| Day 5 | 0.025 |
| Day 6 | 0.017 |
| Day 7 | 0.046 |
| Day 8 | 0.0068 |
| Day 9 | 0.010 |
| Day 10 | 0.0042 |
| Day 11 | 0.00 |
| Day 12 | 0.00 |
| Day 13 | 0.00 |
| Day 14 | 0.0030 |
| Day 15 | 0.0054 |
| Day 16 | 0.00 |
| Day 17 | 0.0014 |
| Day 18 | 0.00 |
| Day 19 | 0.00 |
| Day 20 | 0.00 |
| Day 21 | 0.0030 |
| Day 22 | 0.027 |
| Day 23 | 0.056 |
| Day 24 | 0.026 |
| Day 25 | 0.047 |
| Day 26 | 0.054 |
| Day 27 | 0.077 |
| Day 28 | 0.094 |
| Day 29 | 0.11 |
| Day 30 | 0.15 |
| **Mean FMI** | **0.036** |

**Supplemental Table 1:** Fraction of missing information (FMI) after multiple imputation (MI) via predictive mean matching (PMM). Mean FMI shown at the end of the table.
